# Low frequency of community-acquired bacterial co-infection in patients hospitalized for COVID-19 based on clinical, radiological and microbiological criteria; a retrospective cohort study

**DOI:** 10.1101/2021.06.23.21259020

**Authors:** Sophie Coenen, Jara R. de la Court, David T.P. Buis, Lilian J. Meijboom, Rogier P. Schade, Caroline E. Visser, Reinier van Hest, Marianne Kuijvenhoven, Jan M. Prins, Suzan F.M. Nijman, Elske Sieswerda, Kim C.E. Sigaloff

## Abstract

**Background:** To define the frequency of respiratory community-acquired bacterial co-infection in patients with coronavirus disease 2019 (COVID-19) based on a complete clinical assessment, including prior antibiotic use, clinical characteristics, inflammatory markers, chest computed tomography (CT) results and microbiological test results.

**Methods:** This study was conducted within a cohort of prospectively included patients admitted for COVID-19 in our tertiary medical centres between 1-3-2020 and 1-6-2020. A multidisciplinary study team developed a diagnostic protocol to retrospectively categorize patients as unlikely, possible or probable bacterial co-infection based on clinical, radiological and microbiological parameters in the first 72 hours of admission. Within the three categories, we summarized patient characteristics and antibiotic consumption.

**Results:** Among 281 included COVID-19 patients, bacterial co-infection was classified as unlikely in 233 patients (82.9%), possible in 35 patients (12.4%) and probable in 3 patients (1.1%). Ten patients (3.6%) could not be classified due to inconclusive data. Within 72 hours of hospital admission, 81% of the total study population and 78% of patients classified as unlikely bacterial co-infection received antibiotics.

**Conclusions:** COVID-19 patients are unlikely to have a respiratory community-acquired bacterial co-infection. Prospective studies should define the safety of restrictive antibiotic use in COVID-19 patients.

## Background

Antibiotics are frequently prescribed to COVID-19 patients upon admission out of fear of a bacterial co-infection (1-3). The decision to administer antibiotics is likely influenced by the frequent presence of consolidations on chest computed tomography (CT), high inflammatory markers and frequent bacterial co-infections in patients with influenza (4, 5). Recent studies suggest bacterial co-infection is present in less than 10% of COVID-19 patients (2, 6-8). They seem mostly hospital-acquired. Community-acquired bacterial co-infections occur in 1.2 – 3.5% of patients (6, 7, 9, 10). However, in these studies bacterial co-infection was diagnosed based on microbiological test results only. This could lead to underestimation, since microbiological tests are not obtained in all patients and are negative for the majority of patients with community-acquired pneumonia (CAP) (11). Conversely, positive microbiological test results of respiratory tract samples may represent bacterial colonization.

To date, one study has attempted to assess the prevalence of bacterial CAP in COVID-19 patients, based on clinical, radiological and microbiological criteria (12). The threshold for diagnosing possible bacterial CAP in this study was low, i.e. the presence of one clinical criterion sufficed (such as fever and elevated white blood cell count). The authors reported that in 49% of patients bacterial co-infection was either possible, probable or proven. Given the overlap between symptoms of COVID-19 and bacterial CAP, this percentage may be an overestimation. In order to more reliably identify patients in whom a bacterial CAP is likely, complete patient evaluation of initial presentation and clinical course is essential. We therefore defined the frequency of community-acquired respiratory bacterial co-infection (further abbreviated to co-infection) in COVID-19 patients, based on a complete clinical assessment, including prior antibiotic use, clinical characteristics, inflammatory markers, chest CT and microbiological test results.

## Methods

We performed a retrospective analysis within the CovidPredict Clinical Course Cohort, performed at the two centers of Amsterdam UMC, a tertiary care hospital (13). Data for this cohort was prospectively gathered from electronic health records according to the case report form (CRF) designed by the World Health Organization (WHO, (14)). This study was approved by the Medical Ethics Committee VUmc (Amsterdam, the Netherlands); All necessary patient consent has been obtained and archived.

The cohort includes all adults admitted to Amsterdam UMC with positive SARS-CoV-2 PCR of a respiratory tract sample, or chest CT typical for COVID-19 (COVID-19 Reporting and Data System (CO-RADS) ≥ 4 (15)) since February 2020 (13, 16). Collected data comprises demographics, comorbidities, medication, laboratory tests, microbiological test and chest CT results, treatment and clinical outcomes. Clinical outcome, defined as mortality, admission to ICU or general ward or discharged to their home or rehabilitation centre, is ascertained for this study at 3 weeks after hospital admission by chart review. For the current study, we included patients admitted directly to the Amsterdam UMC from 27-2-2020 to 1-6-2020. This inclusion period represents the first ‘wave’ of COVID-19 in The Netherlands. Exclusion criteria were: nosocomial COVID-19 infection, re-admission or transfer from another hospital or health care institution and no chest CT within 72 hours of admission.

Clinical characteristics included were prior antibiotic use, symptoms at admission (cough, chest pain, dyspnea and confusion), CURB-65 score and inflammatory parameters, i.e. C-reactive protein (CRP) and procalcitonin (PCT). We assessed the clinical course within 72 hours of admission, including admission to ICU, clinical recovery or deterioration, in- or decrease of inflammatory markers in the presence or absence of antibiotics and clinical state in case of hospital discharge.

For the chest CT results we systematically reviewed all reports for the presence of consolidations. Chest CTs without consolidations or only consolidations which were multifocal, crescent-shaped or had round and/or oval morphology were classified not consistent with co-infection. Reports describing large, lobar or unilateral consolidations were reassessed by a chest-radiologist (LM) for definitive classification as consistent or not consistent with co-infection.

Microbiological tests performed within 72 hours of admission were considered. Relevant tests included blood and sputum cultures, bronchoalveolar lavage (BAL) and tracheal fluids, PCRs for *Mycoplasma pneumoniae, Legionella pneumophila* and *Chlamydia pneumonia*, and *Legionella and pneumococcal* urinary antigen tests. Bacteria were considered contaminants when reported as such by the microbiologist.

We categorized patients as unlikely, possible or probable co-infection in two steps. First, patients without respiratory symptoms and without altered mental status, and patients with CRP below 100 mg/L in combination with chest CT not consistent with co-infection and negative microbiological test results were categorized as unlikely co-infection. Second, for all remaining patients categorization was performed by our multidisciplinary study team, hereafter named ‘expert panel’, based on individual patients’ clinical, radiological and microbiological parameters. The expert panel consisted of a chest-radiologist (LM), pulmonologist (SN), infectious disease specialist (KS) and clinical microbiologist (ES). The expert panel used diagnostic criteria to categorize patients as unlikely, possible or probable co-infection (**Additional file 1**: Supplemental tables). First, independent assessments were made by two physicians. Any discrepancies in categorization were resolved in a total of four expert panel meetings.

### Analysis

We performed a descriptive analysis of clinical and laboratory tests. Continuous variables were summarized by mean and standard deviation or by medians and interquartile range (IQR). Categorical variables were presented by counts and percentages. Frequency data were summarized as proportions.

We summarized proportions of patients receiving antibiotics and described the median duration of antibiotic therapy. Overall in-hospital antibiotic use was measured as days of therapy (DOTs), standardized per 100 patient days and per 100 admissions. Patient days were calculated by subtracting day of admission from day of discharge.

Data analysis was performed using R Statistical Software (version 3.6.1; R Foundation for Statistical Computing, Vienna, Austria). Data visualization was performed using TIBCO^®^ Spotfire^®^.

## Results

During the study period, 384 COVID-19 patients were included in the CovidPredict database. We excluded 70 patients who were re-admitted, transferred from another hospital or had nosocomial COVID-19 and 33 patients without chest CT within 72 hours of hospital admission (**Figure 1**). Mean age was 61 years, median CRP 86 mg/L (n=270), median PCT 0.14 ng/mL (n=86, **table 1**). In 43 (15.3%) patients antibiotics were prescribed prior to admission. Three weeks after hospital admission 214 (76%) patients were discharged to their home, a nursing home or rehabilitation unit, 17 (6.0%) were still admitted to the ICU, 15 (5.3%) were still admitted to the general ward and 35 (12%) patients had died or were transferred to palliative care (data not shown).

**Table 1.** Baseline characteristics of the study population.

|  | Probable co-infection<br>(N=3) | Possible co-infection<br>(N= 35) | Unlikely co-infection<br>(N=233) | Total<br>(N=281) |
| --- | --- | --- | --- | --- |
| Male | 2 (66.7%) | 24 (68.6%) | 124 (53.2%) | 157 (55.9%) |
| Female | 1 (33.3%) | 11 (31.4%) | 109 (46.8%) | 124 (44.1%) |
| Age, years |  |  |  |  |
| Mean (SD) | 65.3 (7.8) | 60.1 (13.9) | 60.9 (15.0) | 61.1 (14.7) |
| % ≥ 65 | 33.3 | 34.2 | 41.6 | 42.0 |
| CURB-65 |  |  |  |  |
| 0 | 1 (33.3%) | 9 (25.7%) | 73 (31.3%) | 86 (30.6%) |
| 1 | 1 (33.3%) | 12 (34.3%) | 85 (36.5%) | 99 (35.2%) |
| 2 | 0 (0%) | 8 (22.9%) | 43 (18.5%) | 53 (18.9%) |
| 3 | 1 (33.3%) | 6 (17.1%) | 26 (11.2%) | 35 (12.5%) |
| 4 | 0 (0%) | 0 (0%) | 6 (2.6%) | 8 (2.8%) |
| 5 | 0 (0%) | 0 (0%) | 0 (0%) | 0 (0%) |
| History of hypertension | 0 (0%) | 15 (42.9%) | 107 (45.9%) | 129 (45.9%) |
| Missing | 0 (0%) | 0 (0%) | 2 (0.9%) | 2 (0.7%) |
| History of asthma or COPD | 1 (33.3%) | 9 (25.7%) | 51 (21.9%) | 65 (23.1%) |
| History of chronic kidney disease | 0 (0%) | 6 (17.1%) | 17 (7.3%) | 24 (8.5%) |
| Missing | 0 (0%) | 0 (0%) | 1 (0.4%) | 1 (0.4%) |
| History of liver disease | 0 (0%) | 1 (2.9%) | 12 (5.2%) | 14 (5.0%) |
| History of chronic neurological disorder | 0 (0%) | 3 (8.6%) | 36 (15.5%) | 40 (14.2%) |
| Missing | 0 (0%) | 0 (0%) | 2 (0.9%) | 2 (0.7%) |
| Immunocompromised * | 0 (0%) | 6 (17.1%) | 17 (7.3%) | 24 (8.5%) |
| CRP at admission, mg/L |  |  |  |  |
| Median (IQR) | 110.0 (67.5) | 167.0 (104.5) | 74.0 (86.0) | 85.6 (96.6) |
| Missing | 0 (0%) | 2 (5.7%) | 8 (3.4%) | 11 (3.9%) |
| CRP < 100 mg/L | 1 (33.3%) | 6 (17.1%) | 148 (63.5%) | 155 (55.2%) |
| Missing | 0 (0%) | 2 (5.7%) | 8 (3.4%) | 11 (3.9%) |
| Procalcitonin at admission, ng/ml |  |  |  |  |
| Median (IQR) | 0.27 (3.18) | 0.13 (0.10) | 0.13 (0.24) | 0.14 (0.24) |
| % ≥ 0.25 | 66.7 | 5.7 | 10.3 | 10.7 |
| % ≥ 0.5 | 33.3 | 2.9 | 4.3 | 5.0 |
| Missing | 0 (0%) | 26 (74.3%) | 163 (70.0%) | 195 (69.4%) |
| Antibiotic use prior to admission | 1 (33.3%) | 7 (20.0%) | 31 (13.3%) | 43 15.3%) |

Numbers are n (%) unless otherwise indicated. In 10 patients classification (as probable, possible and unlikely co-infection) was not possible due to inconclusive data.
*\* Immunocompromised is defined as the use of chemotherapy for cancer, bone marrow or organ transplantation, immune deficiencies, poorly controlled HIV or AIDS, or prolonged use of*
corticosteroids or other immunosuppressive medications. CRP: C-reactive protein (CRP).

**Figure 1.**
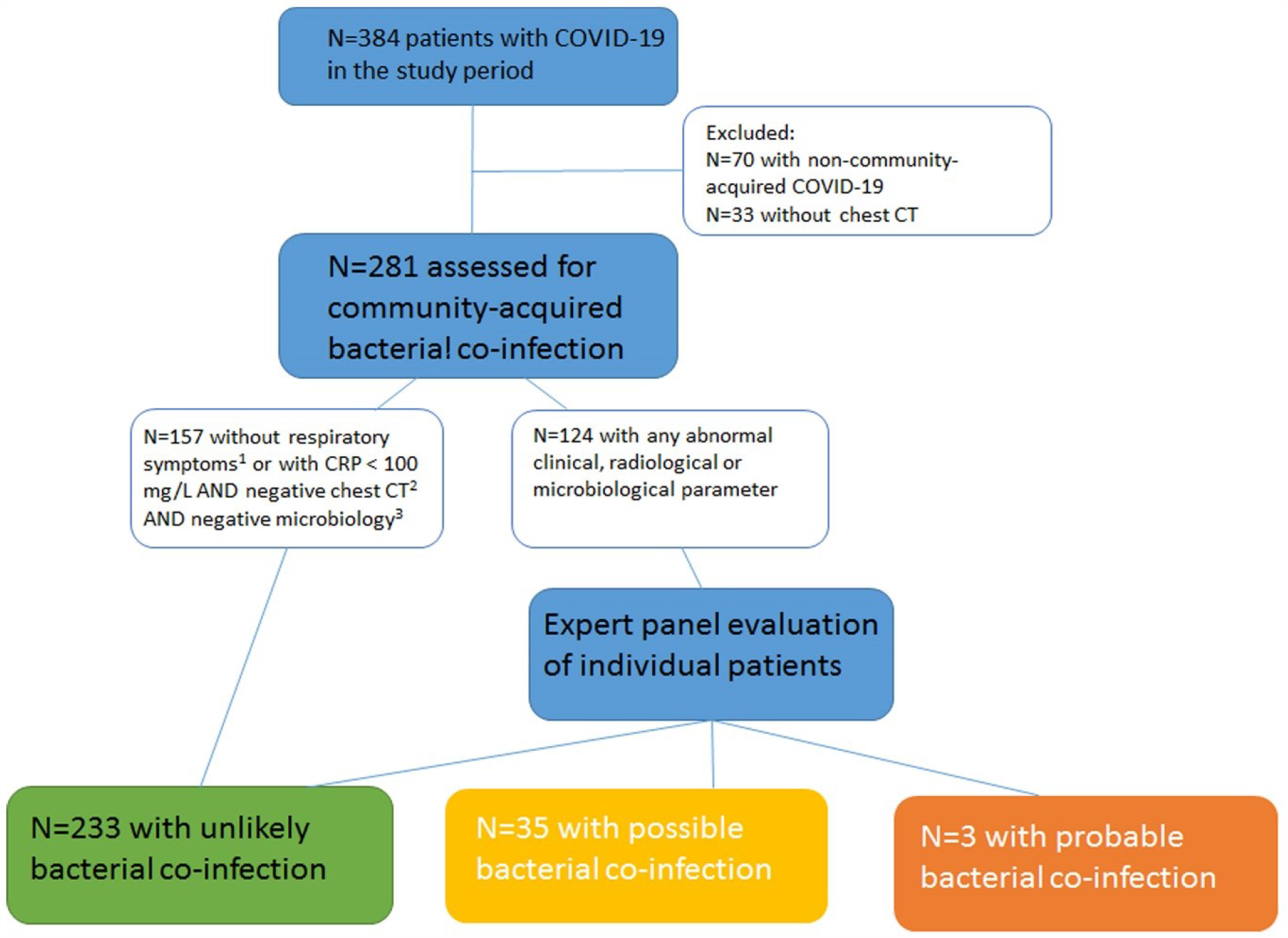
Study flow chart. In 10 patients categorization was not possible due to inconclusive data. ^1^No respiratory symptoms and no altered mental status. ^2^No large, lobar or unilateral consolidations. ^3^Culture, PCR or urinary antigen tests. CT: Computed tomography scan; CRP: C-reactive protein

All included patients underwent chest CT; examples of findings are shown in **Figure 2**. In 104 (37%) patients only ground glass opacity was described. In 166 (59%) patients consolidations were present but not consistent with co-infection. In 11 (4%) patients consolidations were consistent with co-infection.

**Figure 2.**
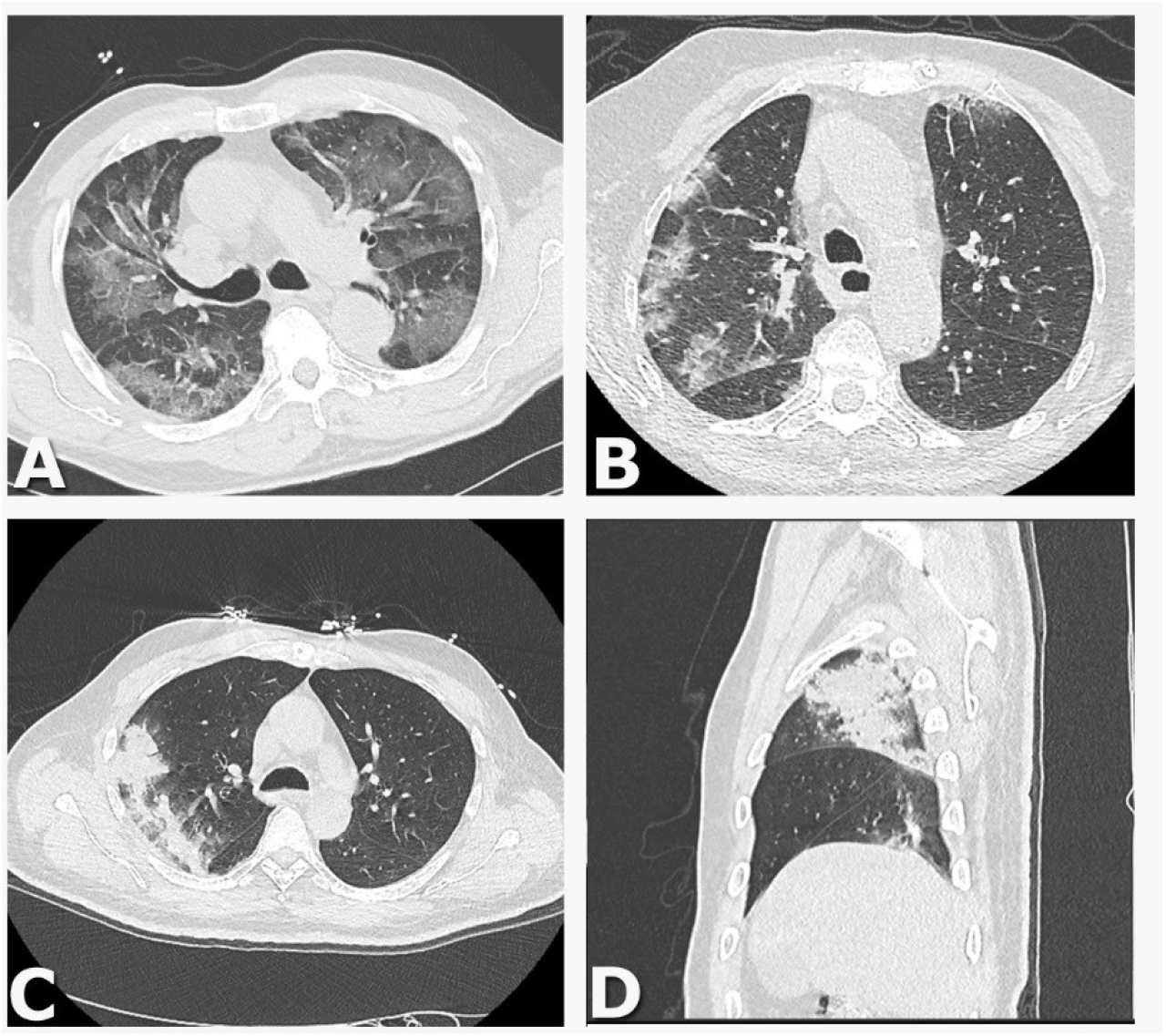
Examples of chest CT findings. A. Axial Chest CT image show multiple areas of pure ground-glass opacity (GGO). B. Axial Chest CT image show peribronchovascular and subpleural patchy consolidations with GGO. C/D. Axial (C) and sagittal (D) Chest CT image show a large consolidation in the right upper lobe with air bronchograms.

In 251 (89%) patients any microbiological diagnostics was performed within 72 hours of admission; in 15 patients (6%) test results were positive (**Additional file 2**: Supplemental tables). Blood cultures were obtained in 232 (83%) patients and yielded positive results in 2 (1%). In 88 (31%) patients sputum cultures were obtained, of which 9 (10%) yielded positive results.

After assessment by the expert panel, co-infection was probable in 3 (1.1%), possible in 35 (12.4%) and unlikely in 233 patients (82.9%, **Figure 1** and **Table 1)**. In 10 (3.6%) patients categorization was not possible due to inconclusive data, i.e. no follow-up inflammatory parameters were available enabling evaluation of the effect of administering or withholding antibiotics.

Of 3 patients classified as probable co-infection, one patient had a PCR-proven *Mycoplasma pneumoniae* infection, with ground glass opacities on chest CT but no consolidations consistent with co-infection. The other two patients did have consolidations consistent with co-infection and one of the patients had a positive urinary pneumococcal antigen test. In all three patients clinical course was consistent with co-infection, i.e. inflammatory markers were elevated and decreased after initiation of antibiotics.

In 7/35 patients classified as possible co-infection, chest CT showed consolidations consistent with co-infection. However, there were no clinical and microbiological findings to support co-infection. In 9/35 patients, microbiological test results were positive but no radiological or clinical findings supported co-infection. In the remaining 19 patients, only clinical findings were consistent with co-infection, i.e. inflammatory markers were markedly elevated and decreased after initiation of antibiotics.

In the 233 patients classified as unlikely co-infection, 33 (14%) had no respiratory symptoms and no altered mental status. In 124 (53%) patients CRP was below 100 mg/L, chest CT was not consistent with co-infection and microbiological test results were negative. In 76 (33%) patients, co-infection was considered unlikely after expert panel review. Of these, in 32 (42%) patients no consolidations were described on chest CT. In 42 (55%) patients, consolidations were present but not consistent with co-infection, and there were no clinical or microbiological findings to support co-infection. Two (3%) patients had unilateral or lobar consolidations on chest CT, but inflammatory parameters were low and the patients’ clinical course improved without initiation of antibiotics. Microbiological test results were positive in 4 (5%) patients classified as unlikely co-infection after expert panel review. In one patient *Staphylococcus aureus* in sputum was considered colonization because there were no clinical or radiological signs of infection. Other positive microbiological test results were due to extra-pulmonary infection (*Escherichia coli* bacteraemia in a patient with urosepsis) or contamination (*Acinetobacter Iwoffii* in blood, *Burkholderia cenocepacia* in BAL).

### Antimicrobial consumption

Antibiotics were initiated in 228 (81%) patients within 72 hours of admission (**Table 2**). Among patients classified as unlikely co-infection, 182 (78%) received antibiotics for a median duration of 4 days (IQR 2-5). In 109 (47%) patients antibiotic treatment was continued beyond 3 days. Among patients classified as possible or probable co-infection, 36 (95%) received antibiotics for a median duration of 5 days (IQR 4-6). Ceftriaxone was the most frequent administered systemic antibiotic with a total of 774 DOT, resulting in 275 DOT ceftriaxone/100 admissions (**Figure 3**). According to the Dutch national guideline on CAP (17), based on the CURB-65 score ceftriaxone could be replaced by amoxicillin in 143/167 (85.6%) patients.

**Table 2.** Antibiotic therapy administered within 72 hours of admission.

|  | Total study population | Co-infection unlikely | Co-infection possible/probable |
| --- | --- | --- | --- |
| Total number of patients | 281 (100) | 233 (100) | 38 (100) |
| Patients receiving systemic antibiotic therapy | 228 (81.1) | 182 (78.1) | 36 (94.7) |
| Ceftriaxone/cefotaxime monotherapy* | 167 (59.4) | 136 (58.4) | 24 (63.2) |
| Ceftriaxone/cefotaxime + quinolone | 25 (8.9) | 17 (7.3) | 5 (13.2) |
| Other combination therapy** | 24 (8.5) | 19 (8.2) | 5 (13.2) |
| Amoxicillin monotherapy | 6 (2.1) | 4 (1.7) | 2 (5.3) |
| Other monotherapy*** | 6 (2.1) | 6 (2.6) | 0 (0) |

Numbers are n (%)
\*Including patients in whom both amoxicillin + ceftriaxone was given.

**Figure 3.**
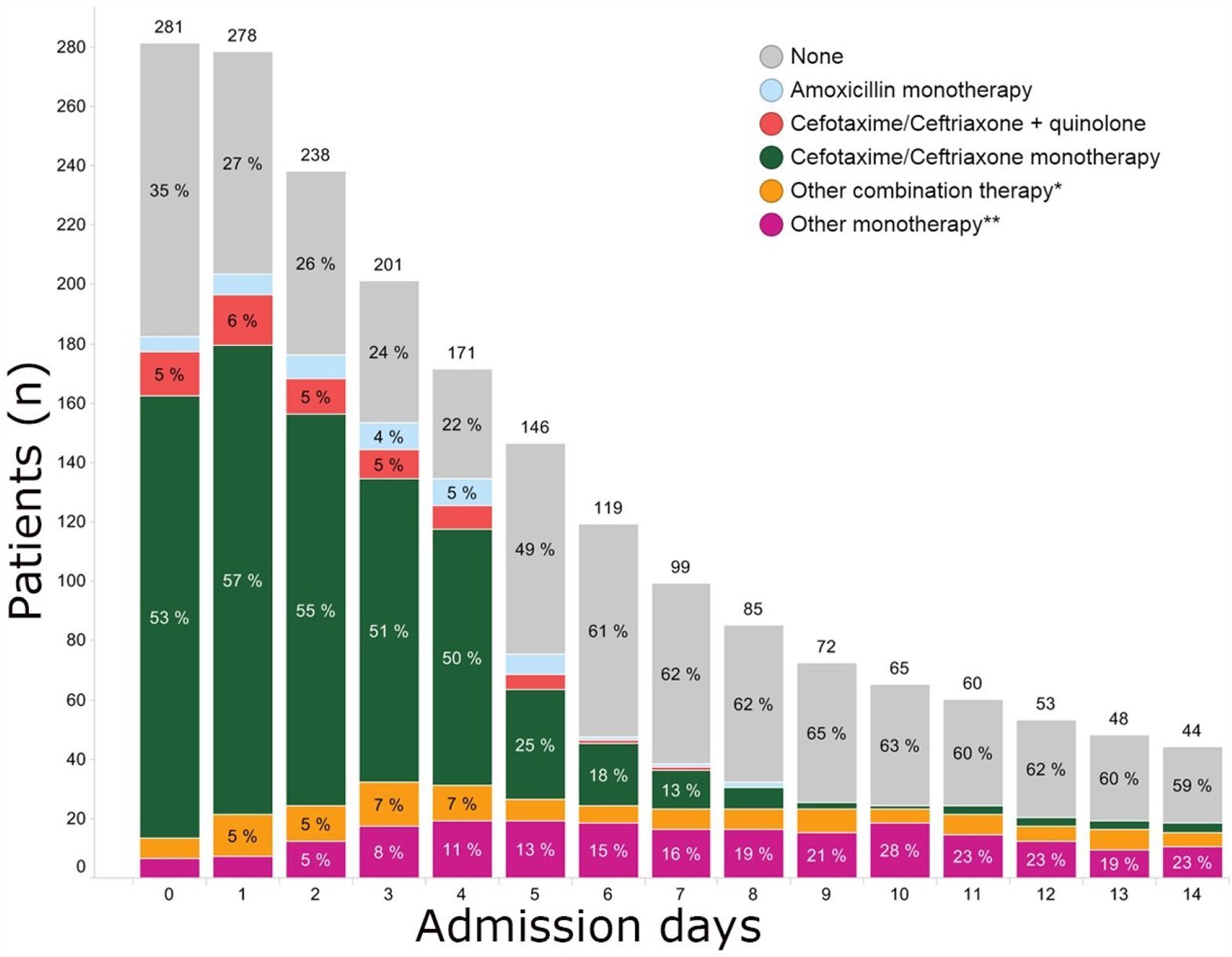
Antibiotic use of total study population per admission day. **\*** Other combination therapy: a combination of beta-lactam antibiotics, combination of beta-lactam antibiotic with macrolide/quinolone/glycopeptide/cotrimoxazole/metronidazole or combinations of three different antibiotic classes. **\*\*** Other monotherapy: other beta-lactam antibiotics (e.g. flucloxacillin or ceftazidime) or any of the other antibiotic classes (e.g. nitrofurantoin, cotrimoxazole).

## Discussion

In our study, 83% of COVID-19 patients had no co-infection based on complete clinical assessment, including prior antibiotic use, clinical characteristics, inflammatory markers, chest CT results and microbiological test results. Using a multidisciplinary approach, we considered co-infection probable in 1.1% and possible in 12.4%. Despite the low frequency of co-infection, antibiotic treatment was initiated in 81% of patients, and ceftriaxone was prescribed most frequently. Our study underpins recent preliminary recommendations for restrictive use of antibacterial drugs in patients with proven or a high likelihood of COVID-19 (18).

Our reported low probably of co-infection is in line with previous study results on co-infections based on microbiological test results only (6, 7, 9, 10). By means of this study, we addressed the limitation of previous reports, i.e. co-infection is not confirmed or ruled out by microbiological test results alone (19). Indeed, we found that only 11/38 (29%) patients classified as possible or probable co-infection had positive microbiological test results. Conversely, 4/15 (27%) positive microbiological test results were considered contamination, colonization or were due to extra-pulmonary infection.

One recent study by Karaba et al. also included clinical and radiological criteria to assess the frequency of co-infection and concluded that co-infection was proven in 0.3%, probable in 1.1% and possible in 48% of patients (12). In contrast to our study, the authors found co-infection unlikely in only 51% of patients. This large difference could be explained by the different criteria used for co-infection. Karaba et al. classified patients meeting one clinical criterion (with the exception of hypoxia) or patients with positive radiological criteria on chest radiograph or chest CT as possible co-infection. This approach is likely to lead to overestimation of possible co-infection. We combined multiple clinical criteria with radiological and microbiological findings and a final classification by an expert panel. Furthermore, Karaba et al. included both chest radiograph and chest CT while chest CT has a higher sensitivity and specificity for bacterial CAP compared to chest radiograph (20).

Chest CT was an important factor in our categorization. No previous studies defined the role of chest CT for co-infections in COVID-19 patients. In patients with compatible clinical characteristics, we considered large, lobar or unilateral consolidations consistent with co-infection, in accordance with international guidelines (21). Chest CT was consistent with co-infection in 9/38 (24%) patients classified as probable or possible co-infection. However, the majority of chest CTs showed typical findings for COVID-19, i.e. ground glass opacities, characteristically with a peripheral and subpleural distribution as described in the CO-RADS scale (15). Such opacities may be mixed with areas of focal consolidation, of which linear consolidations and reversed halo sign suggest organizing pneumonia (22-24).

Strengths of this study include the structured classification of patients, using a predefined, standardized and reproducible clinical evaluation. We reviewed all chest CTs, microbiology data and clinical parameters for each patient. These clinical parameters were prospectively collected and had little missing data. Independent assessments were made by two physicians and discrepancies were resolved by the expert panel. Patients were included prospectively, reducing the risk of selection bias.

Our study has limitations that should be acknowledged. First, our categorization of patients was retrospective in nature which may have biased our categorization. However, we standardized the criteria we used for the final classification to minimize this bias. Second, complete diagnostic work-up was not available in all patients, for example, procalcitonin was only measured in 86 patients.

However, overall there was little missing data as clinical parameters were prospectively collected. Third, atypical bacterial pneumonia may be difficult to distinguish from COVID-19 on chest CT (22-26). Therefore, the frequency of atypical bacterial pneumonia may have been underestimated. However, as only 1 (1.7%) of the 60 tests for atypical pathogens was positive, this risk of bias is deemed low.

Fourth, use of antibiotics prior to admission may have confounded our results. Fifteen percent of patients reported antibiotic use prior to admission, which could have led to underestimation of the frequency of co-infection as prior antibiotic use can lead to false-negative microbiological test results.

Lastly, we studied COVID-19 patients in the first ‘wave’ when patients did not receive corticosteroids yet. The current standard use of corticosteroids for the treatment of COVID-19 may impact the frequency of bacterial infections in this population (27). However, as COVID-19 patients currently only receive corticosteroids when hospitalized, this will most likely not have a large effect on the prevalence of co-infection.

## Conclusions

Our study suggests that implementation of the recent recommendations for restrictive antibacterial therapy in COVID-19 patients has the potential to greatly reduce antibiotics use (18). Although the effect of the pandemic on antimicrobial resistance development is yet unclear, the unnecessary use of antibiotics should be avoided. Up to now, the majority of patients have received broad-spectrum antibiotics which, in combination with prolonged hospital admission, has exposed them to the risk of health-care-associated infections and transmission of (multidrug-)resistant organisms (28, 29).

In conclusion, COVID-19 patients are unlikely to have a co-infection. Withholding antibiotic treatment could lead to a large reduction in unnecessary antibiotic use. Our results underpin the recommendations for restrictive use of antibiotics in recent guidelines. Prospective studies should evaluate clinical course in COVID-19 patients when withholding antibiotics at admission.

## Data Availability

Both Sophie Coenen and Jara R. de la Court have full access to the data used for this manuscript. The data that support the findings of this study are available from the corresponding author, upon reasonable request.

## ABBREVIATIONS

(COVID-19): coronavirus disease 2019
(CT): computed tomography
(CAP): community-acquired pneumonia
(CRF): case report form
(WHO): World Health Organization
(CO-RADS): COVID-19 Reporting and Data System
(CRP): C-reactive protein
(BAL): bronchoalveolar lavage
(IQR): interquartile range
(DOTs): days of therapy
(PCT): procalcitonin

## DECLARATIONS

### Ethics approval and consent to participate

This study was approved by the Medical Ethics Committee VUmc (Amsterdam, the Netherlands); All necessary patient consent has been obtained and archived.

### Consent for publication

Not applicable.

### Authors’ contributions

All authors contributed to the study conception and design. Data collection was performed by Sophie Coenen, data analysis was performed by Jara R. de la Court. The expert panel consisted of Kim C.E. Sigaloff, Elske Sieswerda, Suzan F.M. Nijman and Lilian J. Meijboom, expert meetings were coordinated and documented by Sophie Coenen. The first draft of the manuscript was written by Sophie Coenen and Jara R. de la Court and all authors commented on earlier versions of the manuscript. All authors read and approved the final manuscript.

## Acknowledgments

We would like to thank the CovidPredict consortium (www.covidpredict.org), specifically Joost Wiersinga and Michiel Schinkel, for their efforts in providing the patient data. We also thank the microbiology data-team of the Amsterdam UMC for providing the microbiological data.

