## Supplemental Tables 1 and 2 for "Low frequency of community-acquired bacterial co-infection in patients hospitalized for COVID-19 based on clinical, radiological and microbiological criteria; a retrospective cohort study"

**Supplemental Table 1.** **Parameters used by the expert panel to categorize patients as having unlikely, possible or probable community-acquired bacterial co-infection**

|  | **Co-infection less likely** | **Co-infection more likely** |
| --- | --- | --- |
| Clinical parameters | - CRP < 100 mg/L; or  - No CRP decrease after initiation of antibiotics; or  - Satisfactory clinical response in absence of or <3 days of antibiotics. | -CRP decrease after initiation of antibiotics; or  -Clinical deterioration in absence of antibiotics. |
| Radiology | -Chest CT without consolidations (ground glass opacities only), or;  -Chest CT with consolidations not consistent with bacterial infection^1^. | -Large, lobar or unilateral consolidations on chest CT. |
| Microbiology | -Negative test results^2^; or  -Positive test results due to contamination, colonization or extra-pulmonary infection. | -Positive test results not due to contamination, colonization or extra-pulmonary infection. |

Patients without any respiratory symptoms and without altered mental status, and patients with C-reactive protein (CRP) below 100 mg/L in combination with chest CT findings not consistent with co-infection and negative microbiological test results were categorized as unlikely co-infection. For all remaining patients categorization was performed by the expert panel based on individual patients‘ clinical, radiological and microbiological findings.

^1^Chest CT reports that described consolidations which were multifocal, crescent-shaped or had round and/or oval morphology were classified as not consistent with co-infection. ^2^Culture, PCR or urinary antigen tests.

CT: Computed tomography (CT) scan; CRP: C-reactive protein

**Supplemental Table 2**. **Microbiological tests and identified bacterial pathogens within 72h of admission**

| Diagnostic test | Performed ≤72h, | Positive result | Identified micro-organisms |
| --- | --- | --- | --- |
| Sputum culture | 88 (31) | 9/88 (10) | *7 x Staphylococcus aureus* |
|  |  |  | *1 x Proteus mirabilis* |
|  |  |  | *1 x Haemophilus influenzae* |
|  |  |  | *1 x Moraxella catarrhalis* |
|  |  |  | *1 x Klebsiella pneumoniae* |
| Blood culture* | 232 (83) | 2/232 (1) | *1 x Acinetobacter lwoffi* |
|  |  |  | *1 x Escherichia coli* |
| Broncho-alveolar lavage culture | 4 (1) | 1/4 (25) | *1 x Burkholderia cenocepacia* |
| Urinary pneumococcal antigen test | 104 (37) | 2/104 (2) | *2 x Streptococcus pneumoniae* |
| Atypical pathogens - Urinary legionella antigen test - Mycoplasma pneumonia PCR  - Chlamydia pneumoniae and Chlamydia psittaci PCR | 60 (21) 35 (12)  30 (11)  29 (10) | 1/60 (1.7) 0/35 (0)   1/30 (3) 0/29 (0) | 1 x *Mycoplasma pneumoniae* |
| Any microbiological test performed | 251/281 (89) | 15/251 (6) |  |

Numbers are n(%)
*Coagulase-negative staphylococci were considered contaminants if regarded as contaminants in the clinical consultation of the microbiology staff.
